# Common and Specific Intrinsic Functional Network Related to Episode Dynamics during Treatment in Bipolar Spectrum

**DOI:** 10.1101/2024.04.28.24306505

**Authors:** Xiaobo Liu, Zhen-Qi Liu, Bin Wan, Lang Liu, Jinming Xiao, Yao Meng, Chao Weng, Yujun Gao

## Abstract

A significant challenge in bipolar disorder (BD) is to understand the neural substrates of emotional fluctuations (i.e., episode phases) along the spectrum including manic (BipM), depressive (BipD), and remission states (rBD). Here, We constructed intrinsic functional connectome for 117 subjects with BD (BipM: 38, BipD: 42, and rBD: 37) and 35 healthy controls, then associated connectivities with emotional fluctuations to identify the common and specific patterns, and finally probed their biological underpinnings. We uncovered the common altered pattern in the salience-attention network and the specific pattern in the default mode-salience network specific for BipM and sensory-prefrontal network specific for BipD and rBD. These pathological patterns can accurately delineate the various episodes episodes types of bipolar disorder and forecast the corresponding clinical symptoms associated with each episodes type. Both common and specific patterns exhibited significant genetic stability and centered regions were enriched in multiple receptors such as MOR, NMDA, and H3 for specific pathology while A4B2, 5HTT, and 5HT1a for common pathology. Gene expression was enriched in PLEKHO1, SCN2A, POU3F2, and ANK3. Our study provides new insights into possible neurobiological interpretation for episode phases in the bipolar spectrum and holds promise for advancing personalized precision medicine approaches targeting various episodes of the condition.

## INTRODUCTION

Bipolar disorder (BD) is a psychiatric condition characterized by a spectrum of emotional fluctuations including manic episodes (BipM), depressive episodes (BipD), mixed episodes, and remitted bipolar disorder (rBD). BD affects approximately 2% of the global population, exhibiting high levels of disability, suicide rates, and prevalence[18]. Patients with BD suffer from different functional impairments in various episodes episodes types. For instance, cognitive impairments are linked more closely with BipM rather than BipD or general mood fluctuations [45]. Udergoing therapy in BD, patients experience emotional fluctuations in different states [33]. Understanding the neural substrates related to emotional fluctuations helps strategize treatment for patients with bipolar spectrum.

Previous functional magnetic resonance imaging (fMRI) studies have shown the brain functions of individuals with BD at various episodes states, with a particular focus on changes in resting-state networks and the behavior of differentiated brain regions [37]. These studies primarily concentrated on key brain networks such as the Default Mode Network (DMN), the Central Executive Network (CEN), and emotional regulation networks, revealing significant differences between individuals with BD [53, 56]. By observing reduced or abnormally increased activity in the DMN, study has unveiled issues related to self-reflection and emotional regulation disorders, which manifest differently during the manic and depressive phases of BD [57]. Furthermore, dysfunctions in the CEN have revealed deficits in attention, decision-making, and working memory, associated with cognitive impairments in individuals with BD [47]. Abnormalities in emotional regulation networks are directly linked to problems with emotional processing and emotional stability [11]. Together, resting-state fMRI studies have provided important perspectives for understanding the neural basis of different spisodes episodes type of BD via non-invasively assessing the functional connectivity of pathological regions.

However, neurobiological factors such as gene expression and neurotransmitter systems related to the common and specific patterns have been rarely reported. During manic periods, there may be an increase in dopamine levels, while depressive periods may be associated with a decrease in dopamine levels[4]. Variations in neurotransmitter levels could result in diverse activation patterns within brain networks. Heritability and reliability in functional connectome vary across regions and/or components[46]. Furthermore, gene expression is associated with different neural circuits, subsequently influencing macro-scale brain connectivity patterns. Whereas, the specific and common disease mechanisms between different episodes types of bipolar affective disorder remain unknown. Understanding these differences and connections will not only be beneficial for explaining the possible biological underpinnings of the bipolar spectrum, such as more refined brain connectivity patterns, but also guides subsequent treatments based on psychological medicine, transcranial magnetic stimulation (TMS), or transcranial direct current stimulation (tDCS), advancing research towards precision medicine. Simultaneously, it will also contribute to elucidating the more detailed comorbid mechanisms between different mood disorders and the bipolar spectrum.

Therefore, we hypothesize that there are common and specific network patterns related to emotional fluctuations in the bipolar spectrum. These network patterns are presumed to have a basis in receptors and gene expression. To validate this hypothesis, we construct resting-state intrinsic functional connectome in individuals with three bipolar emotional episodes states and healthy controls, then categorize those pathological edges into common and specific network patterns. Furthermore, to clarify potential neurobiological processing, the common and specific patterns are enriched in the maps of heritability, gene expression, and neurotransmitters and receptors.

## RESULTS

### Common and specific network patterns

We initially presented the average functional connectivity matrices during the remission, manic, and depressive phases of bipolar disorder and compared the statistical difference between patients and healthy controls. Subsequently, we decompose common and specific network patterns within different network modes. We identified that during the manic phase, the nodes exhibiting the greatest network differences are the DorsalFCC of Default Mode Network (DMN), Prefrontal Cortex (PFC), PFCdPFCm, PHC, and Orbitofrontal Cortex (OFC) of Limbic network. In contrast, during the depressive phase, the nodes with the largest network pattern differences are pCun of the Frontoparietal Control Network (FCN), Visual network, and Somatomotor network. Specific patterns in the bipolar remission phase primarily involve OFC in the Limbic network, Par and Cing regions in the FCN, and the Visual network. Shared network patterns across these illness phases are predominantly associated with OFC in the Limbic network, the Dorsal Attention Network (DAN) in the Somatomotor network, and the Ventral Attention Network (VAN). In all, we found that the specific pathological functional networks of BipM were DMN and Limbic, the specific pathological functional networks of BipD were visual and somatomotor network and FCN, and the specific patterns of rBD were FCN and visual network. The shared network patterns of these three episodes types of bipolar disorder were in the Limbic network, DAN, and VAN.

### Validation using machine learning

To validate the network patterns, we implement a machine learning approach to classify patients and healthy controls via both common and specific patterns. We found that the accuracy of the specific pathological network of BipD was 80.56% (with an AUC of 0.87, Pnull < 0.05). The recognition rates of this pattern in BipM and rBD were 59.74% (with an AUC of 0.68) and 56.16% (with an AUC of 0.61), respectively. The accuracy of the specific pathological network of BipM was 71.43% (with an AUC of 0.79, Pnull < 0.05), with recognition rates in BipD and rBD of 51.39% (with an AUC of 0.62) and 52.05% (with an AUC of 0.64), respectively. The accuracy of the specific pathological network of rBD was 75.34% (with an AUC of 0.80, Pnull < 0.05), with recognition rates in BipD and BipM of 51.39% (with an AUC of 0.50) and 58.44% (with an AUC of 0.64), respectively. Shared patterns can be used to identify all mood episodes types, with accuracies of 75% (with an AUC of 0.84) in BipD, 72.73% (with an AUC of 0.78) in BipM, and 79.45% (with an AUC of 0.86) in rBD, respectively.

**TABLE 1.** Demography.

|  | rBD | BipD | BipM | HC |
| --- | --- | --- | --- | --- |
| Age | 24.3±6.5 | 21.9±5.2 | 24.9±6.4 | 26.7±7.3 |
| Sex | 17/20 | 15/27 | 21/17 | 14/21 |
| HRSD | 6.5±2.7 | 22.6±3.3 | 5.6±0.8 |  |
| YMRS | 50.2±9.8 | 52.2±10.0 | 52.8±12.1 |  |
| PDQ | 34.7±9.0 | 45.4±13.3 | 31.5±12.2 |  |
| HAMA | 12.2±4.8 | 24.9±7.8 | 17.9±7.0 |  |

HRSD means, YMRS means, PDQ means, HAMA means

### Biological stability of the significant patterns

To test the biological stability of the functional edges, we analyzed the test-retest reliability and heritability in the twin-designed Human Functional connectome (HCP). Reliability reflects intra-individual robustness over time [31] and heritability reflects inter-individual biological robustness [41]. We assume that the stability and heritability of the lesion network pattern in the originally healthy population are high, which aids in further elucidating whether the pathogenic mechanism of the disease involves changes in epigenetic inheritance or phenotypic inheritance, resulting in unstable network connections [54].Briefly, we calculated the realibity (intraclass correlation coefficient, ICC) and narrow-sense heritability (*h*_2_) for the whole connectome edges.

We found 75% of specific pattern edges in BipM, 100% in BipD, and 95% in rBD were significantly stable, and average ICC were 0.37 *±* 0.14, 0.59 *±* 0.09, and 0.48 *±* 0.14 respectively. Regarding the common network pattern, we found that 85.71% of edges were significantly reliable, and the average ICC was 0.41 *±* 0.12.

In addition, we found that the average heritability of the specific pattern was 0.17 ± 0.089 in rBD, 0.31 ± 0.071 in BipD, and 0.27 ± 0.075 in BioM. While the average heritability of the common pattern was 0.22 ± 0.066. Moreover, the proportion of significant edges (p < 0.05) to the specific pattern was 53.49% in rBD, 100% in BipD, and 94.44% in BipM. However, only 14.30% of the common pattern showed statistical significance.

### Static and dynamic structure of specific and common network pattern and clinical symptom

In the context of rBD, our investigation revealed noteworthy correlations within specific pathological hubs. Specifically, a statistically significant positive correlation was identified between degree and HAMA score in the rBD pathological hub (*R* = 0.28, *P* = 0.002). Similarly, in the BipM pathological hub, there was a significantly positive correlation between degree and YMRS score (*R* = 0.37, *P* < 0.001), while in the shared pathological hub, a positive correlation was found between degree and PDQ scores (*R* = 0.24, *P* = 0.009). Additionally, within the BipD pathological hub, a significant negative relationship emerged between the HRSD score and average degree (*R* = −0.23, *P* = 0.012).

Moving on to the assessment of average switch rates within both specific network patterns and shared network patterns alongside cognitive factors, intriguing patterns emerged. Notably, a significant negative association was observed between the HRSD score and the average switch rate in the BipD pathological hub (*R* = −0.25, *P* = 0.005). Similarly, in the BipM pathological hub, a negative correlation was found between the YMRS score and average switch rate (*R* = −0.30, *P* = 0.001), and in the shared pathological hub, a negative relationship was identified between the PDQ score and average switch rate (*R* = −0.30, *P* = 0.0011). Additionally, a positive relationship surfaced between the HAMA score and the average switch rate in rBD hubs (*R* = 0.30, *P* = 0.001).

### Structural basis of common pathological networks

We conducted an in-depth investigation into common pathological nodes characterized by similar structural foundations. Subsequently, we undertook a comparative analysis of cortical thickness within the default mode network across diverse episodes types of specific diseases. Our findings revealed a notable increase in cortical thickness in individuals with rBD compared to HC (*t*= 2.77,*P* = 0.007). Conversely, BipM manifested a significantly diminished cortical thickness relative to HC (*t* = 2.77,*P* = 0.007). Notably, no statistically significant specificion in cortical thickness emerged between individuals with BipD and the HC group (*t* = 0.87, *P* = 0.390). Moreover, the cortical thickness of rBD exhibited no significant disparity compared to BipD (*t* = 1.83, *P* = 0.071) but demonstrated a significant elevation compared to BipM (*t* = 4.72, *P* < 0.001). Likewise, BipD exhibited a significantly higher cortical thickness compared to BipM (*t* = 3.01, *P* = 0.004).

### Molecular mechanisms of specific and common network patterns

Finally, we conducted a thorough analysis of the molecular and genetic foundations associated with specific and shared patterns during different phases of bipolar disorder. Notably, during the remission phase, receptors including 5HT1a, 5HT1b, 5HT4, 5HTT, A4B2, D1, D2, DAT, NET, and NMDA were found to be significantly linked to specific networks (*P*_*F DR*_, *P*_*spin*_ < 0.05). Similarly, in the depressive phase, specific networks (*P*_*F DR*_, *P*_*spin*_ < 0.05) were associated with receptors such as 5HT1a, 5HT4, 5HTT, A4B2, CB1, D1, D2, DAT, MOR, and NET. Networks specific to the manic phase (*P*_*F DR*_, *P*_*spin*_ < 0.05) were related to receptors, including 5HT1a, 5HT1b, 5HT2a, 5HT4, 5HTT, A4B2, D1, D2, DAT, H3, and NET. Furthermore, receptors associated with shared network patterns (*P*_*F DR*_, *P*_*spin*_ < 0.05) across phases included 5HT1a, 5HT1b, 5HT4, 5HTT, A4B2, D2, DAT, H3, and NET. Additionally, our findings revealed that 5HT1a, 5HT4, 5HTT, A4B2, D1, D2, DAT and NET are implicated in both specific and common networks across different phases of bipolar disorder. NMDA was significantly related to rBD, MOR was significantly related to BipD, and H3 was significantly related to BipM.

On the genetic level, during the remission phase, specific network patterns were associated (*P*_*F DR*_, *P*_*spin*_ < 0.05) with PLEKHO1, SCN2A, POU3F2, and ANK3. Conversely, the manic phase (*P*_*F DR*_, *P*_*spin*_ < 0.05) exhibited specific network patterns associated with SCN2A, POU3F2, and ANK3. Common network patterns were identified with PLEKHO1, LMAN2L, SCN2A, and POU3F2 (*P*_*F DR*_, *P*_*spin*_ < 0.05). Notably, no specific network patterns related to the depressive phase were found to be associated with genes implicated in bipolar disorder.

## DISCUSSION

In the present study, using multi-center functional and structural magnetic resonance imaging techniques, we are the first to extract specific and common network patterns associated with various episodes types of Bp. Subsequently, we analyzed these highly stable and genetically influenced network patterns in relation to diverse pathological cognitive and molecular foundations, aiming to assess the specific and shared network relationships with other mood disorders. Our findings revealed that: 1. BipM exhibited specific pathological alterations predominantly within the DMN and limbic network, while BipD displayed aberrations in the somato-motor network and FCN), and rBD showcased disruptions in the FCN and visual network. Common differential patterns were predominantly observed in the limbic network, DAN, and VAN. 2. Both common and specific network patterns exhibited high stability and heritability. 3. Static and dynamic configurations of these network patterns demonstrated significant correlations with clinical symptoms. 4. Common pathological hubs manifested specific structural characteristics, including increased cortical thickness in rBD, decreased cortical thickness in BipM, and no significant differences in cortical thickness in BipD. 5. specific network patterns were predominantly modulated by different receptors and genes. In summary, our study revealed both specific and common pathological networks associated with bipolar affective disorder, providing a theoretical basis for subsequent individualized clinical treatments.

### specific and common patterns in different episodes types of bipolar disorder

Our study was the first one to present novel insights into the specific and common pathological functional networks associated with BipM, BipD, and rBD phases, elucidating their connections with emotional and cognitive domains. Firstly, BipM’s specific pathological networks primarily involve the default mode and limbic networks. Previous studies have shown that the DMN is closely associated with self-awareness, introspection, and emotion regulation [1]. The limbic network plays a crucial role in emotion processing and memory formation [2]. Our research further suggests that during manic episodes, abnormalities in these networks may lead to excessive emotional arousal and disturbances in self-perception, which may be related to manic-specific emotional outbursts and impulsive behaviors. BipD’s specific pathological networks are found in the somatomotor network and FCN. Previous research has indicated that the somatomotor network is involved in motor control [3], while the FCN is associated with cognitive control and emotion regulation. Abnormalities in these networks during depressive episodes may result in slowed movement, decreased mood, and impaired cognitive function, explaining the occurrence of depression-specific clinical symptoms. Importantly, changes in functional connectivity within these two large-scale brain networks may reflect abnormalities in sensory and cognitive integration. rBD’s specific pathological networks are observed in the visual network and FCN, with the visual network being implicated in visual information processing[4], and the FCN playing a critical role in cognitive control [5]. Abnormalities in these networks during bipolar remission may lead to difficulties in processing visual information and cognitive control, suggesting that cognitive dysfunction in rBD patients does not recover with emotional stabilization, indicating persistent abnormalities in brain functional networks. These specific pathological network abnormalities can be interpreted as the biological basis for cognitive impairment at different episodes types and can facilitate the development of personalized treatment strategies to improve treatment outcomes.

Secondly, the common pathological networks of BipM, BipD, and rBD are primarily located in the limbic network, DAN, and VAN. Similarly, previous research has revealed that the limbic network is associated with emotion processing, the DAN is primarily responsible for orientation and sustaining attention, especially controlling external stimuli, and the VAN is related to visual information processing, attentional shifting, and emotion processing. These common pathological networks may reflect the coordinated action of these three key networks across different episodes types of bipolar disorder, and the imbalance in emotion regulation, cognitive control, and attention may be common features and core symptoms of bipolar disorder. These findings underscore the shared neural network abnormalities at different episodes types, providing a theoretical basis for developing more effective treatment strategies by understanding the association between these shared network abnormalities and core symptoms.

### Pathological network properties reveal clinical symptom

Additionally, this study integrates brain network nodal degree with clinical symptoms, and nodal degree is used to describe the connectivity density of brain networks, depicting the role of a node in information transmission and integration across the entire network, which is related to synaptic plasticity and changes in neuronal connectivity patterns [48]. We found that the nodal degree of lesioned brain areas in BipM positively correlates with manic clinical presentations. And increased nodal degree in BipM patients may signify a state of heightened activity, consistent with previous research. These regions become more active, leading to instability in decision-making and increased impulsive behavior [14], possibly reflecting the overactivity of certain brain regions during mania, which is a core symptom of bipolar disorder. Conversely, during depressive episodes, the nodal degree of lesioned brain areas in BipD patients negatively correlates with clinical depressive symptoms, suggesting reduced connectivity density in these brain regions during depression. This aligns with decreased neurotransmitter, such as dopamine[9] and glutamate[6], emphasizing the important role of these specific brain regions in the pathogenesis of depressive disorders. In rBD patients, from the perspective of neuroplasticity and adaptability, these findings may indicate neuroplastic changes in these regions [15], where treatment could lead to the reorganization of neuronal connections to adapt to new environments or situations, potentially correlating with symptom relief in patients. Lastly, the nodal degree of common lesioned brain areas negatively correlates with the degree of cognitive impairment, highlighting the significant impact of these regions on emotion regulation and cognitive function in bipolar disorder patients. These research findings contribute to a more comprehensive understanding of the neurobiological basis of bipolar disorder, providing deeper insights for future treatments.

The switch rate response of brain networks and the spatiotemporal configuration of brain function [7]. The consistency between dynamic and static network attribute results verifies the robustness of network lesions over time scales, jointly confirming the relationship between the information transmission and integration capabilities of these lesioned areas and clinical manifestations from a temporal and overall perspective, further supporting the specificity and commonality of different network patterns in bipolar disorder at different episodes types.

### The structural basis of common functional networks across the three episodes types of bipolar affective disorder is inconsistent

The thickness of the cortex reflects the density and arrangement of neurons in cortical regions of the brain, which in turn affects the organization and coordination of functions among different brain regions[34]. We further showed the common pathological hubs at various disease episodes types revealed different structural foundations and elucidate long-term cortical change. Shared functional connections suggest that the shared connectivity strength across different episodes types of bipolar affective disorder is significantly stronger in the limbic network than in healthy controls, while structural studies show a significant decrease in cortical thickness in this region for BipM, a significant increase for rBD, and no difference for BipD. There results align with previous findings [19, 59]. The results of functional connectivity first explain the limbic network-based compensatory mechanism across different episodes types of bipolar affective disorder, possibly to adapt to changes in patient emotional states and self-regulation of brain function [7]. Enhanced functional connectivity may represent more efficient information transmission or stronger cooperative activities between certain brain regions to maintain normal cognitive and emotional regulatory functions [49]. The differential structural basis of this functional compensatory change may be related to the emotional excitement in BipM, leading to structural damage, i.e., reduced neuronal density, and the structural compensation during remission may be related to the relief of emotional symptoms and self-regulation mechanisms. The functional differences in BipD did not result in changes in cortical thickness, possibly because compensatory regulation during depression is a short-term change [49]. These results provide valuable insights for subsequent clinical treatments, suggesting that targeted treatments at different episodes types may have different side effects.

### The molecular and genetic basis of specific and common pathological networks in bipolar disorder

Firstly, we examine the relationship between brain-specific and common networks across different episodes types of BD and receptors, with all episodes type functional patterns being associated with 5HT receptors. The functional pathological networks of BD primarily involve the default mode network, limbic network, and ventral attention network, associated with emotion regulation and cognitive control, such as attentional control[35, 42]. Serotonin receptors and their subclasses are primarily involved in central nervous system excitatory behavior and mood fluctuations [5], with elevated levels associated with mania [24] and decreased levels associated with depression [30]. Our findings further suggest that 5HT receptors may influence emotion and cognitive imbalance in BD by regulating key pathological nodes within the default mode and limbic networks and the connectivity patterns throughout the brain. Similarly, we find that A4B2 is associated with all specific and shared network patterns. This receptor in ventral attention network may involve the regulation of attentional networks under acetylcholine modulation, impacting functions related to cognition, perception processing, and attention allocation [2, 38]. Dopamine receptors D1, D2, DAT, and NET may induce connectivity abnormalities in the default mode network, limbic network, and ventral attention network across different episodes types through dopamine level regulation and interactions with other neurotransmitter systems, affecting brain network function, including emotion and cognitive control, and thus contributing to the pathological processes of the disease [13, 25, 50]. The rBD is typically a relatively stable and recovery period during the course of the disease [23]. During this episodes type, neuroadaptation may occur as the brain attempts to adapt to previous abnormalities. NMDA receptors may play a role in neuroadaptation and recovery processes, particularly in lesions involving visual, motor sensory networks, and higher-order cognitive networks (such as FCN) [21]. NMDA receptors are closely related to the glutamate system, an excitatory neurotransmitter. During the remission episodes type of BD, the excitatory neurotransmitter system may undergo a balancing state, influencing the function of visual, motor sensory networks, and higher-order cognitive networks [58].

Additionally, BD is a complex psychiatric disorder involving the interaction of multiple genetic factors. Genes identified in the above studies, such as PLEKHO1, SCN2A, POU3F2, and ANK3, may play significant roles in the genetic basis of the disease [44]. These genes may affect emotion regulation and cognitive function by influencing neuron function, synaptic transmission, and neural plasticity pathways [27]. The relevance of genes such as PLEKHO1, SCN2A, POU3F2, and ANK3 during the remission episodes type suggests their involvement in disease state regulation ikeda2018genome,smeland2020genetic. These genes may participate in regulating neuron homeostasis and neurotransmission, affecting the neural function of patients during the remission period. The correlations between the manic phase pattern and SCN2A, POU3F2, and ANK3, and the involvement of PLEKHO1, LMAN2L, SCN2A, and POU3F2 in the shared pattern, may reflect the roles of these genes at different episodes types of the disease course [43]. The existence of shared patterns may indicate that these genes have a certain influence throughout the course of the disease, while specific patterns may reflect their relative importance at different episodes types of the disease course. The lack of specific genetic patterns during the depressive phase may indicate that the genetic basis of the depressive phase may be more diverse or influenced by environmental factors[39]. The findings provide clues for the gene-phenotype association in BD, aiding in understanding the biological basis of the disease. This may pave the way for the development of more precise treatment methods, such as targeted gene therapy or pharmacological interventions.

### Limitation

Noteworthy are certain limitations in this study. Firstly, the sample for this research is derived from a single center, and the sample size is relatively small, potentially constraining the reliability and generalizability of the results. Future endeavors plan to conduct multicenter studies with larger samples to address the limitations associated with a single center and a relatively small sample size in this study. Simultaneously, efforts will be made to enhance the external validity of the research, thereby increasing result generalizability and facilitating the extrapolation of study findings to a broader patient population. Secondly, during the course of the investigation, this study recruited both initial BipM patients and those with rBD beyond the medication washout period to gain a comprehensive understanding of the imaging features of BD. Given the potential impact of medications on brain structure and function, it is imperative to acknowledge this as a limitation in our study. This may be an unavoidable challenge faced by other studies as well. Presently, an ideal approach to address this issue has yet to be identified. Finally, this study solely employed a longitudinal rating scale to assess symptom changes in BipM patients before and after treatment, while the remainder followed a cross-sectional design. Consequently, the study was unable to observe the trajectory of structural and functional neuroimaging changes in the brains of BD patients. Subsequent research endeavors will contemplate implementing longitudinal follow-up assessments to capture the longitudinal variations in brain structure, function, as well as emotional and cognitive functions across different periods in individuals with BD.

## CONCLUSION

In this study, utilizing multi-center structural and functional magnetic resonance imaging data, for the first time extracted specific pathological networks and common pathological network patterns associated with different episodes types of bipolar affective disorder. Unique network patterns were correlated with clinical symptoms at different episodes types, while common pathological networks not only were associated with cognition but also exhibited different structural bases across different mood episodes types, revealing differences in disease-related functional and structural alterations. The more robust and heritable pathological patterns were found to be underpinned by different receptors and genes. This represents the first comprehensive investigation into the pathological mechanisms of bipolar affective disorder across episodes types, encompassing structural, functional, and receptor aspects, providing a research foundation for comprehensive individualized treatments from brain network TMS to gene targets for subsequent individuals.

## METHODS

### Dataset 1: Bipolar Disorder

#### Clinical symptoms and cognitive assessment

All data collection was approved by the Ethics Committee of Wuhan University People’s Hospital. The Young Mania Rating Scale (YMRS) [13] is a clinical assessment tool specifically designed to evaluate manic symptoms and their severity, developed by Dr. Paul H. Young and colleagues in 1978. It consists of 11 items, with scores ranging from 0 to 4 for items 1, 2, 3, 4, 7, 10, and 11, and from 0 to 8 for items 5, 6, 8, and 9. Severity levels of manic symptoms are determined based on the total score: Normal (0-5), Mild (6-12), Moderate (13-19), Severe (20-29), Very Severe (30 and above). Clinicians assess each item based on patient responses, and the total score guides the understanding and management of manic symptoms.

The Hamilton Anxiety Scale (HAMA) [14] was initially designed by Dr. Max Hamilton in 1959 to objectively and reliably assess anxiety symptoms. The revised 1980 version includes 14 items, scored from 0 to 4, representing levels of symptom severity: (0) None, (1) Mild, (2) Moderate, (3) Severe, (4) Very Severe. Anxiety factors are categorized into somatic and psychic, and scores are analyzed accordingly. Chinese criteria categorize scores as: (1) Total score *≥* 29 indicates severe anxiety; (2) *≥* 21 indicates definite anxiety; (3) *≥* 14 indicates presence of anxiety; (4) *>*7 suggests possible anxiety; (5) *<*7 indicates absence of anxiety symptoms.

The Hamilton Depression Rating Scale (HAMD) is widely used in clinical settings for assessing depression severity. Developed by Max Hamilton in 1960, the HAMD-17 is a newer version indicating disease severity through lower scores for milder conditions and higher scores for more severe cases. HAMD is categorized into seven factors: (1) Anxiety/Somatization, (2) Weight, (3) Cognitive Disturbances, (4) Diurnal Variation, (5) Retardation, (6) Sleep Disturbances, and (7) Desperation. Chinese criteria categorize scores as: (1) Total score <7 indicates normal; (2) 7-17 suggests possible depressive symptoms; (3) 17-24 indicates definite depressive symptoms; (4) >24 indicates severe depressive symptoms. The Perceived Disability Questionnaire (PDQ) assesses the degree of impairment in daily life, work, social, and leisure activities. It consists of 20 items, each scored on a 5-point scale from 0 (no difficulty) to 4 (extremely difficult). The questionnaire covers various functional activities such as concentration, work completion, social interaction, and household chores.

In this study, two trained doctoral students in psychiatry conducted neurocognitive assessments on all participants in the same environment, with each assessment taking approximately 60 minutes.

#### Imaging

We Collected data from patients diagnosed with Bipolar Disorder who sought treatment at the Department of Psychiatry and Psychology, Wuhan University People’s Hospital, from September 2021 to December 2023. Initial screening of research participants was conducted using the Chinese version of the Mini-International Neuropsychiatric Interview (MINI) [3]. All participants were diagnosed by two experienced psychiatrists following the Diagnostic and Statistical Manual of Mental Disorders Fifth Edition (DSM-5) criteria for BD [10]. Inclusion and exclusion criteria for the BipM group: Age 18-45; DSM-5 diagnosis of BD, YMRS > 7, and HAMD < 7; First episode and untreated or first-time undergoing treatment.criteria for BipD group was HRSD-17 > 7 and YMRS < 7.Inclusion criteria for rBD group: Age 18-45; DSM-5 diagnosis of BD, HAMD < 7, and YMRS < 12; Patient-initiated discontinuation for more than 14 days. Exclusion criteria: MRI contraindications; Organic brain diseases; Other mental illnesses; History of medication or physical therapy for BipM patients; Left-handedness; Unstable physical illnesses; Substance abuse history; Pregnancy or lactation; Concurrent other brain function disorders.

HCs were recruited through community, university, and Hubei Provincial People’s Hospital posters, with age and gender to the patient group; right-handedness. Exclusion criteria for HCs: MRI contraindications; Organic brain diseases; Substance abuse history; Pregnancy or lactation; Family history of neurological or psychiatric disorders. All participants had withdrawal and termination criteria: Withdrawal criteria included voluntary withdrawal of informed consent and researcher judgment of unsuitability to continue. Termination criteria involved non-cooperation leading to data invalidation, exclusion from final data analysis, and discontinuation of further investigation.

MRI images were acquired using an Achieva 3T MRI scanner (GE, SIGNA Architect) equipped with a 48-channel headcoil. Participants were instructed to stay awake, remain motionless, relax, and keep their eyes closed during the scanning procedure. To minimize head movement and reduce scan noise, foam padding and soft earplugs were provided. All scans were performed by two licensed MRI technicians with intermediate professional titles in the MRI room. T1_3D data sets were acquired with a maximum TR, minimum TE, a NEX of 1, a layer thickness of 2mm, and a field of view of 256 × 256 mm^2^. Scan time = 7 minutes. For rs-fMRI, a TR of 2000ms, a TE of 30ms, a FOV of 220mm *×* 220mm, a flip angle of 90°, a matrix of 64 *×* 64, a resolution of 3*×* 3 *×* 3, a slice thickness of 36, and 240 time points were acquired. Scan time = 9 minutes.

### Data preprocessing and Functional connectome

For all datasets, raw DICOM files were converted to Brain Imaging Data Structure (BIDS) format [17] using HeuDiConv v0.13.1 [12]. The structural and functional preprocessing of both Bipolar were performed with fMRIPrep 23.0.2 [17], which is based on Nipype 1.8.6 [18]. The main anatomical data preprocessing steps include intensity normalization, brain extraction, tissue segmentation, surface reconstruction, and spatial normalization. The main functional data preprocessing steps include head motion correction, slice-time correction, and co-registration. For original preprocessing details generated by fMRIPrep, see Supplementary X. The derived functional time series were parcellated into the Schae-fer200×7 atlas [19] and underwent a confound removal process implemented in Nilearn [20]. The confound removal process adopted the “simple” strategy from [21], including high-pass filtering, motion and tissue signals removal, detrending, and z-scoring. Functional connectivity matrices were then estimated for each subject using zero-lag Pearson correlation coefficient.

### Common network pattern and specific network pattern

To extract pathological changes of the networks, we first compared the functional connectome between manic patients, depressive patients and euthymic patients with healthy control respectively via two-sample t-test with False Discovery Rate (FDR) with 0.05 threshold. We delineate common and specificive network patterns across diverse disease cohorts through the overlapping and non-overlapping sets of edges within specific differential network profiles. It is noteworthy that specific disease network patterns could only diagnose the specific episodes type, whereas common pathological patterns can identify different disease episodes types and normal individuals. Threfore,to validate the specificity of network patterns, we additionally employed machine learning techniques to ascertain the capacity of these network patterns in discerning variations across different disease cohorts. Specifically, we implemented Support Vector Machine (SVM) with a radial basis kernel and default parameters to classify patients and healthy controls. We split the data into training and testing sets via leave-one-out and selected network patterns using specific and common network profiles. Finally, the model performance was evaluated using accuracy and Area Under Curve (AUC). Additionally, we built a null model for comparison. We randomly selected the same number of edges consistent with specific and common networks in diseases and normal individuals to compose network patterns. We used SVM classification to obtain accuracy and repeated the entire process 1000 times.

### Stability and heritability of functional connectivity

Next, we estimated functional connectivity from the two aspects including test-retest reliability and heritability, which reflect intra-subject robustness across time and biological robustness determined by behavioral genetics. To do so, we utilized the Human Connectome Project (HCP) S1200 data release for large sample size of healthy adult population. [51]. This dataset comprise four sessions of resting-state functional magnetic resonance imaging (rs-fMRI) scans from 1206 healthy young adults, along with pedigree information, including 298 monozygotic and 188 dizygotic twins, as well as 720 singletons. We specifically selected individuals with a complete set of four fMRI scans that met the HCP quality assessment criteria [16, 52]. Ultimately, our sample encompassed 1014 subjects (470 males) with an average age of 28.7 years (range: 22–37) and 45 retest (approximately 140-day interval) subjects (13 males).

We assessed the test-retest reliability of FC by calculating the intra-class correlation coefficient (ICC) [40]. The ICC is defined as 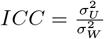, where 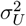 denotes the intra-subject variance and 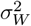 the total variance. Total variance is the sum of intra-subject variance and inter-subject variance. To interpret the results [8], an ICC-score less than 0.40 is poor; between 0.40 and 0.59 is fair; between 0.60 and 0.74 is good; and between 0.75 and 1.00 is excellent.

To map the heritability of functional gradient asymmetry in humans, we used the Sequential Oligogenic Linkage Analysis Routines (SOLAR, v8.5.1b)[1]. In brief, heritability indicates the impact of genetic relatedness on a phenotype of interest. SOLAR uses maximum likelihood variance decomposition methods to determine the relative importance of familial and environmental influences on a phenotype by modeling the covariance among family members as a function of genetic proximity [1, 54]. Heritability (i.e. narrow-sense heritability *h*^2^) represents the proportion of the phenotypic variance 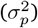 accounted for by the total additive genetic variance 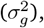, that is 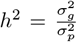. Phenotypes exhibiting stronger covariances between genetically more similar individuals than between genetically less similar individuals have higher heritability. In this study, we quantified the heritability of functional gradients. We added covariates to our models including age, sex, age^2^, and age *×* sex. We calculated heritability in all two sessions and averaged them as final heritability.

### Static and dynamic properties of pathological network patterns

Node degree indicates the extent of a node’s connections with others, serving as a metric for node centrality and reflecting its importance in the network topology. By comprehensively examining node degree, we gain a holistic understanding of the functionality and position of nodes in the diseased brain network. Here, we first extracted the top 10% of connectome matrix, binaried connection matrix and calculated binary-based node degrees via Brain Connectome Toolbox (BCT, https://sites.google.com/site/bctnet/) [36].

The pathology of mental disorder could be represented as the switching of multiple resting networks[29]. The BOLD time sequence was divided into several short segments by sliding window method[22], and the whole brain FC matrix of each segment was calculated. A time window of 50 TR in length and 1 TR in step was used to segment the data. At each window, we calculated the Pearson correlation coefficient between the BOLD time course for each pair of ROIs and obtrained dynamic function connectome (dFC). The dFC matrix obtained was the Fisher’s z-transformed correlation coefficient matrix of all time points, and its dimension is 200 *×* 200 *×* 182, where 200 is the ROI number and 182 is the time number. Then, we then estimated dynamic network switch rate [32] to summarize the transition of each ROIs across time points. Noted that switch rate was based on an iterative and ordinal Louvain algorithm and represented the percentage of time windows when brain nodes transitions among various network assignments [32].

### The clinical characterization of static and dynamic dynamics within network patterns

To explore clinical symptoms of extracted networks of different patients, we further linked dynamic and static properties to clinical questionaires. Specifically, we firstly defined the regions with largest number of significant edges as pathological hubs and then extracted their network properties. Finally, we calculated Pearson’s correlation between switch rate and degree of those pathological hubs in BipM,rBD, BipD and their common pathological regions with YMRS,HRSD,HAMA, and PDQ respectively.

### Structural foundation of functional pathological hubs

To investigate whether the common functional pathological hubs in rBD, BipD, and BipM share similar structural underpinnings, we extracted the cortical thickness data corresponding to those common pathological hubs for each individual. Subsequently, we computed the average cortical thickness values across these shared pathological hubs within network patterns for each patient, thereby simplifying statistical analyses. This approach allowed for an assessment of the overarching structural mechanisms governing these functional pathological hubs. Following this, we conducted two sample t-tests to compare the average cortical thickness among individuals with rBD, BipD, and BipM.

### Precise pathological network patterns extraction

Differential indicators of seed-based functional connectivity (sFC) hold key significance in understanding treatment response in BD patients. The sFC network can establish potential predictive models for treatment outcomes by comprehensively analyzing the strength and patterns of connections between different regions[55]. Fmri was susceptible to environmental noise [23], and exhibited substantial individual variability [24]. Therefore, considering the noise and instability of functional connections, we compared the robustness and heritability of the different regions in the disease in healthy adult in HCP, filtering to get a few edges with high stability and high heritability (p <0.05). Then the number of filtered egdes that each node has was used to define pathological hubs. And pathological hubs top 5 filtered edge number was slected as seed to construct the network connection mode with other brain regions. A representative set of specific and Common cortical patterns were then averaged.

### Receptor maps from Positron Emission Tomography

Receptor densities were assessed through the utilization of PET tracer investigations encompassing a total of 18 receptors and transporters spanning nine neurotransmitter systems. These data, recently shared by Hansen and colleagues [28] (https://github.com/netneurolab/hansen_receptors). The neurotransmitter systems include dopamine (D_1_, D_2_, DAT), norepinephrine (NET), serotonin (5-HT_1A_, 5-HT_1B_, 5-HT_2_, 5-HT_4_, 5-HT_6_, 5-HTT), acetylcholine (*α*_4_*β*_2_, M_1_, VAChT), glutamate (mGluR_5_), GABA (GABA_A_), histamine (H_3_), cannabinoid (CB_1_), and opioid (MOR). Volumetric PET images were aligned with the MNI-ICBM 152 nonlinear 2009 (version c, asymmetric) template. These images, averaged across participants within each study, were subsequently parcellated into Shaeffer 200 template. Receptors/transporters exhibiting more than one mean image of the same tracer (5-HT_1B_, D_2_, VAChT) were amalgamated using a weighted average.

### Genetic expression mechanisms

In order to study how the pathological network patterns is regulated by genes, we combined Allen Human Brain Atlas (AHBA; http://human.brain-map.org/) and the precise brain connectivity pattern for analysis. Regional microarray expression data were obtained from six post-mortem brains provided by the Allen Human Brain Atlas. We used the abagen toolbox(https://github.com/netneurolab/abagen) to process and map the data to 200 parcellated brain regions from Schaefer parcellation. We first extracted the gene expression [44] associated with risking of bipolar disorder from ENIGMA toolbox [26], and finally calculated the correlation between the brain maps of these gene expression and the specific and common connection patterns of different episodes types of bipolar disorder.

### Null model

In our current investigation, a persistent inquiry revolves around the topographic correlation existing between common-specific network patterns and other salient features. In order to draw inferences regarding these correlations, we have employed a null model designed to systematically disrupt the relationship between two topographic maps while preserving their spatial autocorrelation. We firstly shuffled receptormaps according to [20] and calculated the relationship between this map and common-specific network patterns. These resulting spatial coordinates served as the foundation for generating null models through the application of randomly-sampled rotations and the reassignment of node values based on the nearest resulting parcel, a process iterated 1000 times. Notably, the rotation was initially applied to one hemisphere and then mirrored onto the other hemisphere.It is noteworthy that the 95th percentile of shuffling occurrence frequencies derived from both spatial and temporal null models was designated as the threshold value.

## Data Availability

All data produced in the present study are available upon reasonable request to the authors

https://www.researchgate.net/profile/Xiaobo-Liu-15

## DATA AVAILABILITY

The clinical data could be accessed according to reasonalble request for corresponding authers. The raw fMRI data and MRI data for HCP was available on https://db.humanconnectome.org/. Heritability analyses were performed using Solar Eclipse 8.5.1b (https://www.solar-eclipse-genetics.org). Neuromap (https://netneurolab.github.io/neuromaps/usage.html), ENIGMA toolbox (https://enigma-toolbox.readthedocs.io/en/latest/pages.html).

## CODE AVAILABILITY

Code will be available upon reasonable request.

## ACKNOWLEDGMENTS

Xiaobo Liu is supported by China Scholarship Council. Bin Wan is supported by International Max Planck Research School on Neuroscience of Communication: Function, Structure, and Plasticity (IMPRS NeuroCom), Graduate Academy Leipzig, and Mitacs Globalink Research Award.

---

**Figure 1.**
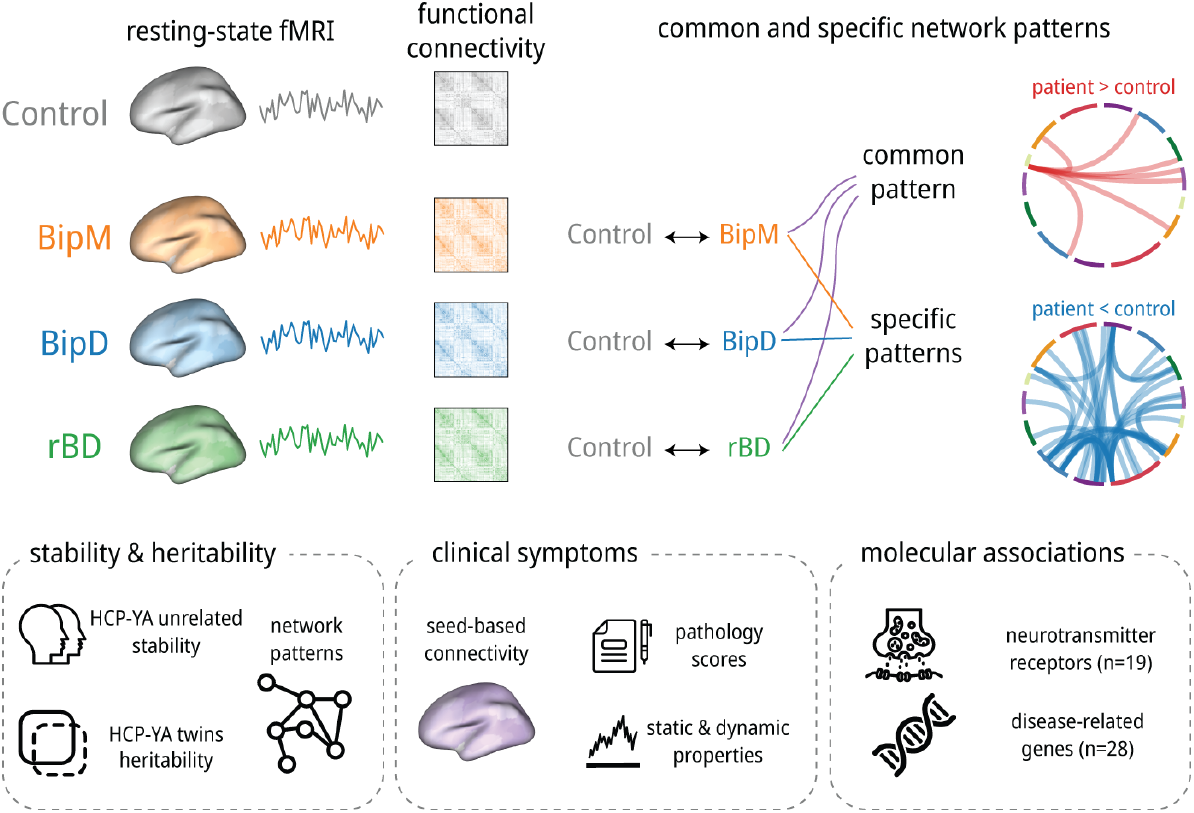
Overview. We first constructed functional connectivity and compared them between different disease episodes types Then we divided those connectivities into common and specific patterns and validated them using machine learning. We further evaluated the test-retest reliability and twin-based heritability of those pathological patterns and linked their properties with clinical symptoms. Finally, we explored the potential molecular mechanisms of those patterns.

**Figure 2.**
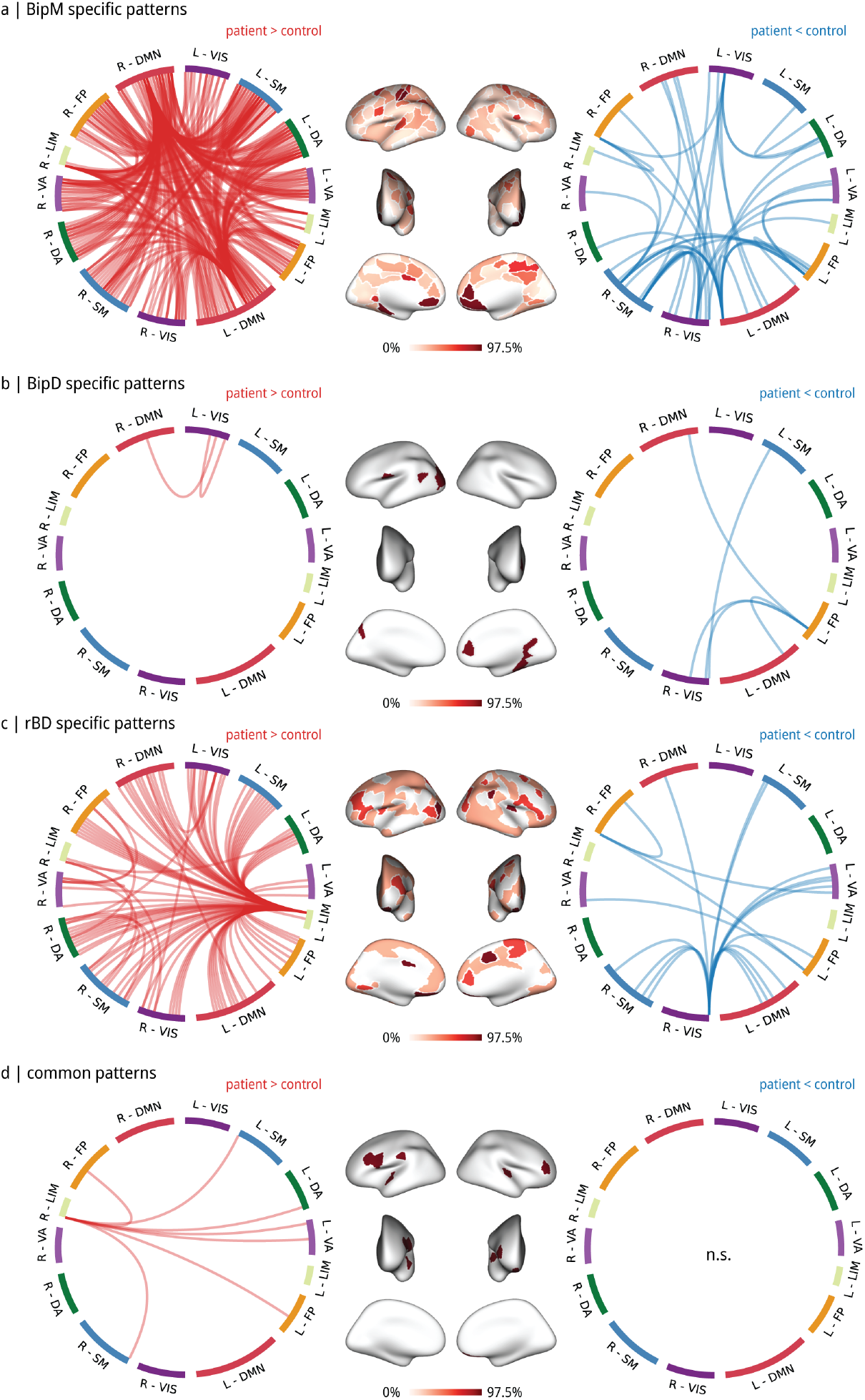
Common and specific network patterns. **(a)** Specific pathogenic network patterns of BipD **(a)**, BipM **(b)**, and rBD **(c). (d)** Shared network patterns. The different color regions in the pie chart represent the proportion of 7 resting-state networks in both specific network group and shared network group. We present average functional connectivity matrices in remission, manic, and depressive phases of bipolar disorder. We then analyze common and specificive patterns in different network modes. In the manic phase, DorsalFCC (Default Mode Network), PFC, PFCdPFCm, PHC, and OFC (Limbic network) nodes exhibit significant network differences. Conversely, in the depressive phase, pCun (Frontoparietal Control Network), Visual network, and Somatomotor network nodes show the largest differences. Bipolar remission phase-specific patterns involve OFC (Limbic network), Par and Cing regions (Frontoparietal Control Network), and Visual network. Shared patterns across phases are linked to OFC (Limbic network), Dorsal attention (Somatomotor network), and SalVentAttation.

**Figure 3.**
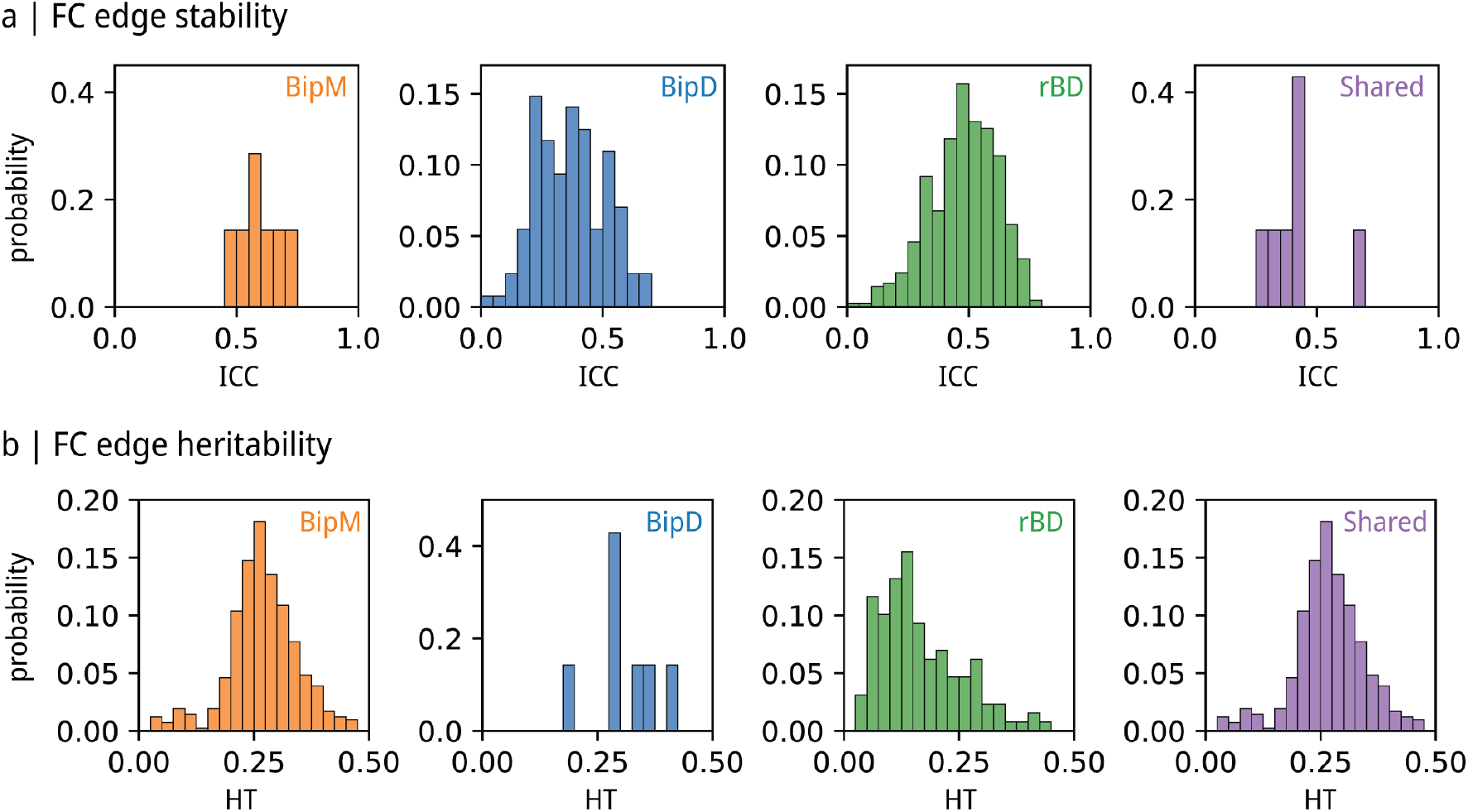
Stability and heritability of Common and specific patterns via HCP test-retest dataset and twins dataset. **(a)** Stability of FCs patterns. Note that the pie chart represents the proportion of significant edges (pICC < 0.05, color portion) and not significant (pICC > 0.05, grey portion) edges within specific pathological edges. **(b)** Heritability of FCs patterns. Noted that the pie chart represents proportion of significant edges (pHeritability < 0.05, color portion) and not significant (pHeritability> 0.05, grey portion) edges within specific pathological edges.

**Figure 4.**
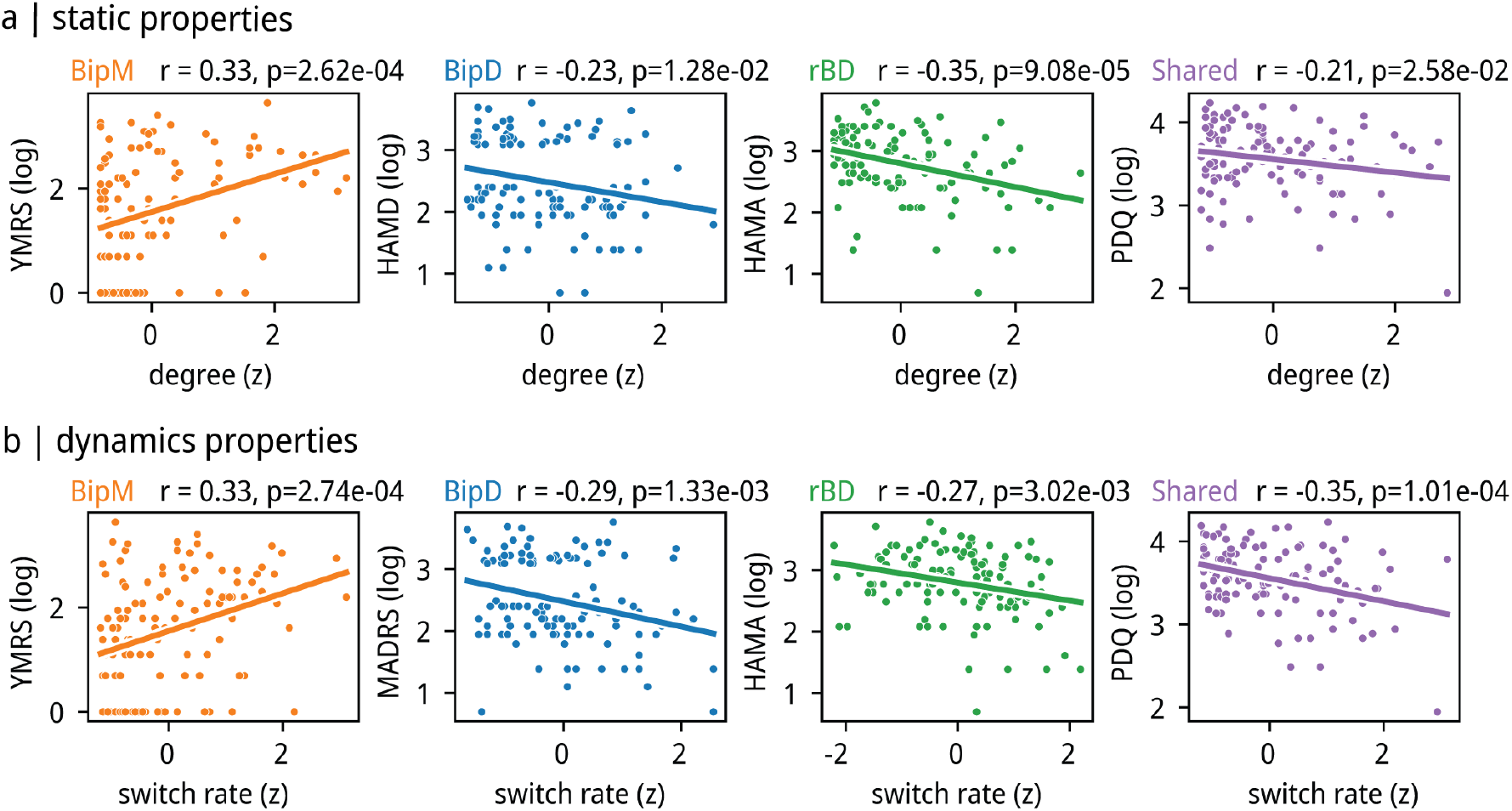
Static and dynamic properties of specific-common pathological hubs. **(a)** The correlation between average degree and cognition in the pathological hubs of both specific network patterns and shared network patterns. Specifically, the observed correlation was significantly negative between degree and HAMA score (p = 0.017, R =- 0.22) in rBD pathological hub, PDQ scores in shared pathological hub (p = 0.019, R = - 0.22) as well as degree and HRSD score and average degree in BipD pathological hub (p = 0.012, R = - 0.23). Also, we found significantly positive relationship between degree and YMRS score (p = 3.94 × 10-5, R = 0.37) in BipM pathological hub. **(b)** Average switch rate in the pathological hubs of both specific network patterns and common network patterns and cognition. We found the significant negative relationship between HRSD score and average switch rate (p = 0.0050, R = −0.26) in BipD pathological hub, average switch rate in shared pathological hub (p = 0.0011, R = - 0.30), PDQ score and HAMA score and average switch rate in rBD hubs (p = 0.0077, R = −0.25). Also, we found positive relationship between YMRS score and average switch rate (p = 0.0036, R = 0.27) in BipM pathological hub.

**Figure 5.**
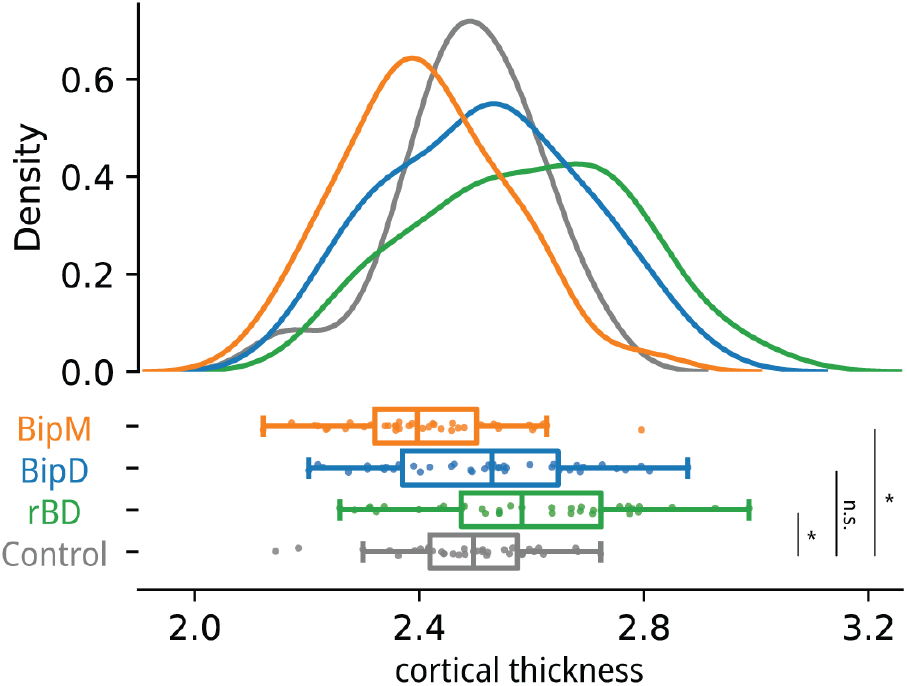
The cortical thickness of rBD, BipD, BipM and HC in common regions. **(a)** We further investigated shared pathological nodes with similar structural foundations; thus, we compared the cortical thickness of the default mode network across different episodes types of specific diseases. We observed a significantly increased cortical thickness in rBD compared to HC (p = 0.0071, t = 2.77), while BipM exhibited a significantly lower cortical thickness than HC (p = 0.0071, t =-2.77). However, no significant difference in cortical thickness was found between BipD and healthy controls (p = 0.39, t = 0.87). Correspondingly, the cortical thickness of rBD showed no significant difference from BipD (p = 0.071, t = 1.83) but was significantly higher than BipM (p = 0.000011, t = 4.72). Similarly, BipD exhibited a significantly higher cortical thickness than BipM (p = 0.0036, t = 3.01).

**Figure 6.**
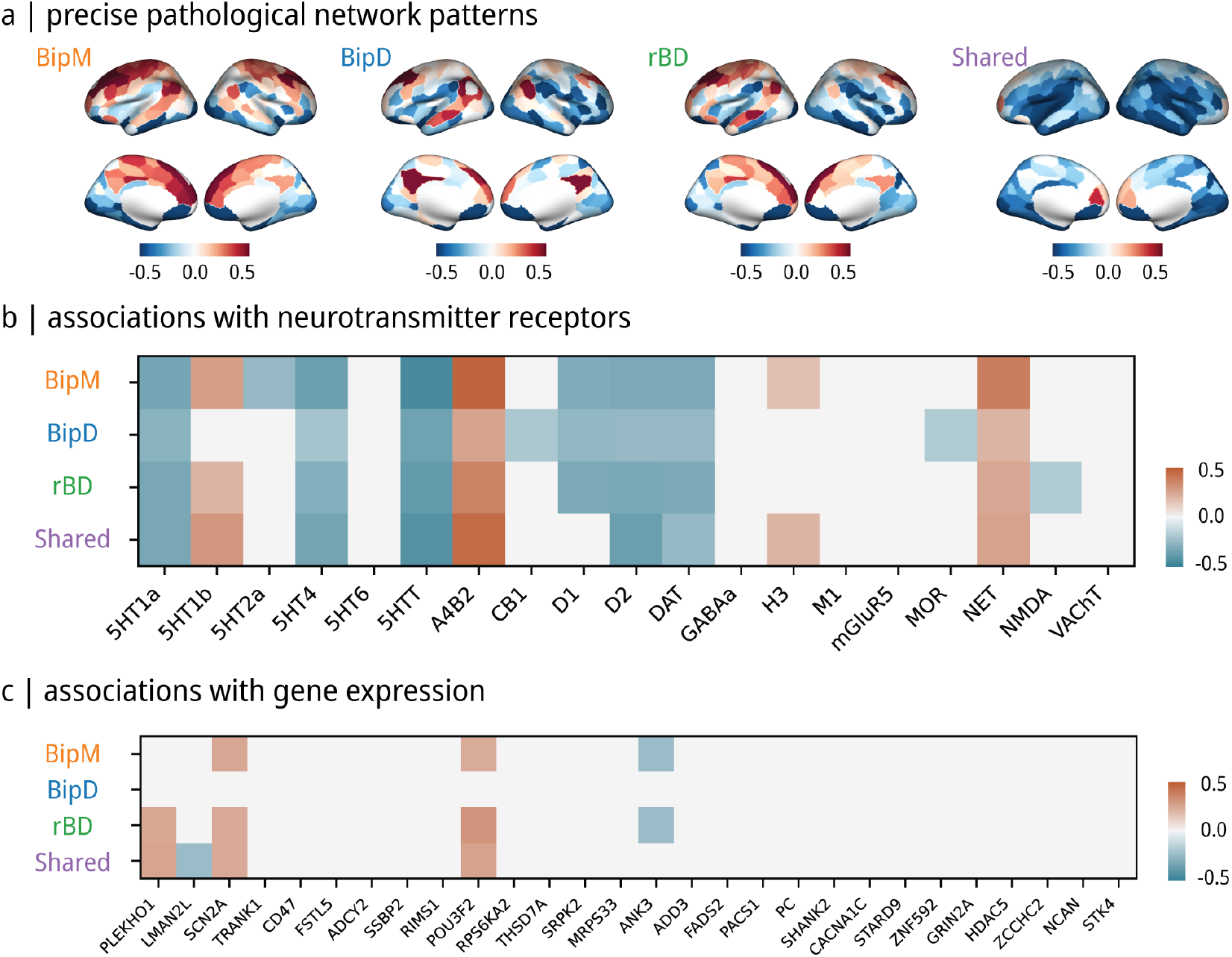
Molecular Structure of specific-Common regions based functional patterns. **(a)** Receptor mechanism of specific network patterns. **(b)** genetic mechanism of specific network patterns. Our study explored molecular and genetic aspects of bipolar disorder phases. During the remission phase, networks were identified that exhibited associations with receptors, including 5HT1a, 5HT1b, 5HT4, 5HTT, A4B2, D1, D2, DAT, NET, and NMDA. In the depressive phase, networks were correlated with receptors such as 5HT1a, 5HT4, 5HTT, A4B2, CB1, D1, D2, DAT, MOR, and NET. Manic phase networks displayed relationships with receptors 5HT1a, 5HT1b, 5HT2a, 5HT4, 5HTT, A4B2, D1, D2, DAT, H3, and NET. Shared networks across phases involved receptors 5HT1a, 5HT1b, 5HT4, 5HTT, A4B2, D2, DAT, H3, and NET. At the genetic level, remission was associated with PLEKHO1, SCN2A, POU3F2, and ANK3. Manic phase patterns were linked to SCN2A, POU3F2, and ANK3, with common patterns involving PLEKHO1, LMAN2L, SCN2A, and POU3F2. No phase-specific genetic patterns were identified for the depressive phase in association with bipolar disorder-related genes.

## Supplemental Materials

